# Ambient air pollution and the sex ratio at birth: a systematic review and narrative synthesis

**DOI:** 10.1101/2025.10.21.25338487

**Authors:** Ursula Gazeley, Hallie Eilerts-Spinelli, Jasmin Abdel Ghany, Ana Bonell, Joshua Wilde

## Abstract

Ambient air pollution is associated with adverse pregnancy outcomes such as stillbirth, miscarriage, and preterm birth. However, its effects on the viability, implantation, and survival of conceptuses—largely unobserved before the clinical detection of pregnancy— remain poorly understood. Shifts in the sex ratio at birth (SRB) provide a sensitive, population-level indicator of sex-biased conception and pregnancy loss. This systematic review examined whether maternal exposure to ambient air pollution before conception and during pregnancy is associated with deviations in the SRB. We searched MEDLINE, Embase, and Global Health from inception to 12 February 2025 for observational studies assessing both ambient air pollution exposure and SRB. Eleven studies met the inclusion criteria, representing data from eight countries spanning 1970–2023. PM_10_ was the most frequently investigated pollutant, followed by PM_2.5_. Six studies reported no significant association, while four observed increased feminisation with higher exposure levels. Air pollution may plausibly influence SRB through effects on gamete quality, conceptus viability, and placental development. The SRB therefore provides a valuable proxy for detecting early biological vulnerability, revealing effects of air pollution that are otherwise hidden due to the selective observation of clinically recognised pregnancies. However, current evidence remains limited, heterogeneous, and inconclusive. As ambient air quality continues to deteriorate, more standardised research is needed to improve comparability and quantify the impact of air pollution on reproductive outcomes.

## 1. Introduction

Ambient air pollution has been consistently associated with adverse pregnancy outcomes, including miscarriage and stillbirth, and adverse perinatal outcomes, such as preterm birth, small for gestational age (SGA), and low birth weight (LBW) (Nyadanu *et al*., 2022; Song *et al*., 2023). These effects are well established in both high-income and low- and middle-income countries (LMICs). However, these observed outcomes represent only a selective subset of pregnancies that survive beyond clinical recognition to later gestational ages. (Wilcox *et al*., 1988; Ammon Avalos, Galindo and Li, 2012). As a result, much of the impact of air pollution exposure on conception, implantation, and early embryonic development remains unmeasured.

Many, if not most, human conceptions do not end in a live birth (Wilcox *et al*., 1988; Ammon Avalos, Galindo and Li, 2012). Early pregnancy losses occurring before clinical recognition are typically unobserved or unrecorded outside of assisted reproduction. These losses can be inferred at the population-level using the sex ratio at birth (SRB) – defined as the ratio of male to female live births (Bruckner and Catalano, 2018; Catalano, Casey and Bruckner, 2020). In the absence of sex-selective induced abortion, the natural human SRB is typically considered to fluctuate around 103-105 males per 100 live born females (Ein-Mor *et al*., 2010; Chao *et al*., 2019). However, evidence suggests that acute or chronic maternal stressors can reduce this ratio, leading to more female live births than expected (Bruckner and Catalano, 2018; Catalano, Casey and Bruckner, 2020). Deviations from the baseline SRB may arise through two, potentially co-occurring pathways: (1) sex-biased conception, which alters the likelihood of conceiving a male versus female zygote; and (2) sex-biased pregnancy loss, including failures at implantation and differential survival of male versus female conceptuses in utero, including miscarriage and stillbirth. Identifying changes in the SRB associated with maternal exposures can therefore serve as a useful proxy for otherwise unobserved changes in reproductive health.

A range of environmental exposures – including extreme heat stress (Abdel Ghany *et al*., 2024), cold stress (Catalano, Bruckner and Smith, 2008), water pollution (Long *et al*., 2021), exposure to toxins such as PCBs, dioxins, pesticides, and lead (Terrell, Hartnett and Marcus, 2011; Azizi *et al*., 2024) – have been associated with reductions in the SRB, resulting in a skew towards female live births. Biological mechanisms remain unclear, but endocrine disrupting toxins may affect the sex ratio at conception via the production, viability, or motility of Y-bearing sperm (James, 2001). After conception, male conceptuses may be more susceptible to environmental stressors due to faster growth rates, higher metabolic demands, and less resilient placental function, making them more vulnerable under adverse conditions (Clifton, 2010; Saoi *et al*., 2020). Consistent with the Trivers-Willard hypothesis (Trivers and Willard, 1973)— that female offspring have higher reproductive success than males when maternal condition is poor—male-biased vulnerability in utero offers a plausible evolutionary explanation for observed SRB shifts.

While the independent effects of air pollution on pregnancy outcomes, and of acute maternal stressors on the SRB, are relatively well documented, it remains unclear whether air pollution exposure contributes to shifts in the SRB at the population-level (Terrell, Hartnett and Marcus, 2011). Conceptually, several plausible pathways through which air pollution could affect the SRB are illustrated in **Figure 1**. To date, however, no systematic review has focused exclusively on ambient air pollution and the SRB. The only prior review considered environmental pollutants broadly, with air pollutants as a subset, and is now outdated (Azizi *et al*., 2024). A targeted, up-to-date synthesis is necessary to assess the specific contribution of air pollution to the SRB.

**Figure 1.**
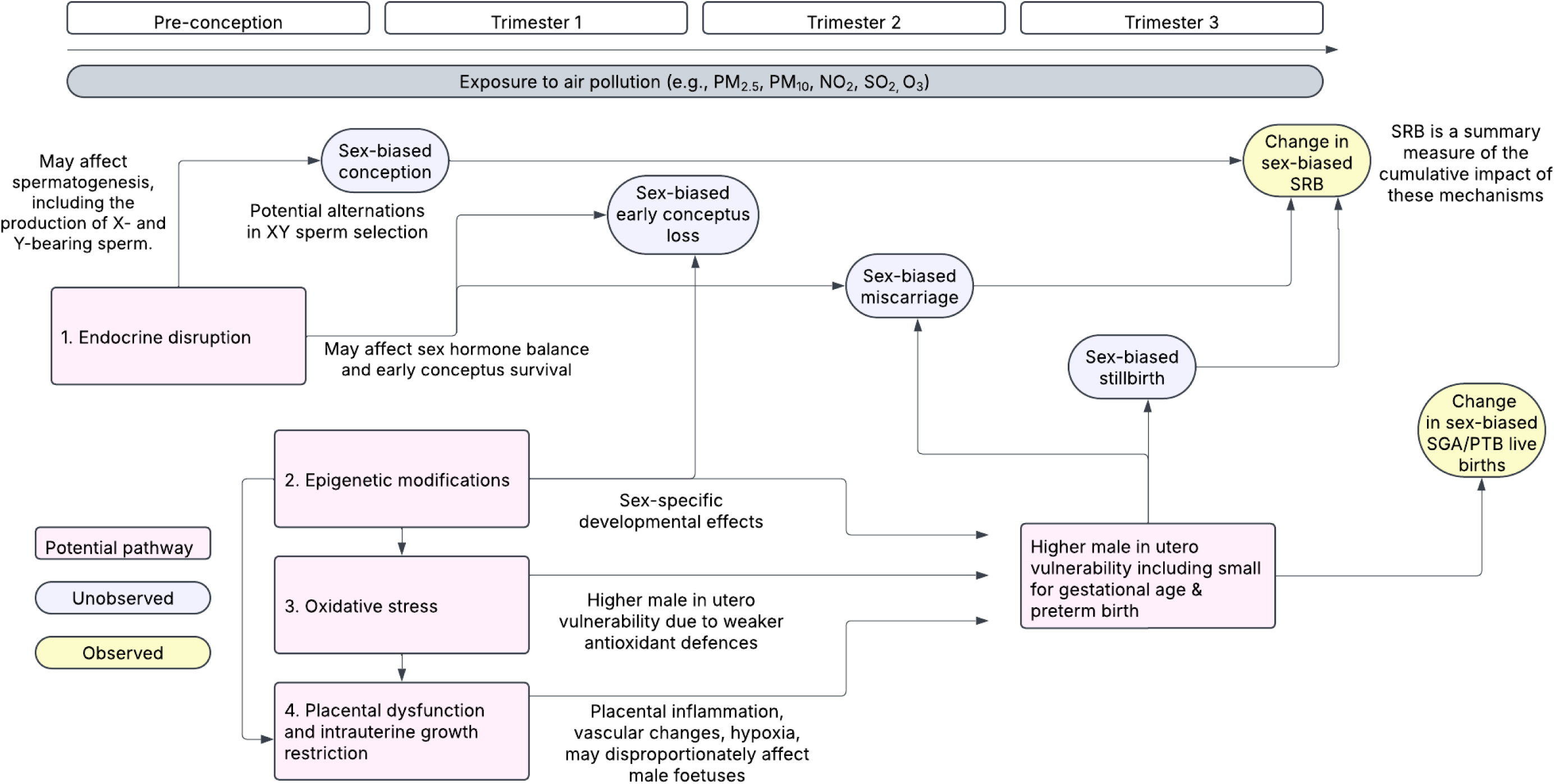
Conceptual framework of potential mechanisms of air pollution exposure on the sex ratio at birth.

Change in the SRB could serve as a sentinel marker of reproductive health (Bruckner and Catalano, 2018; James and Grech, 2018), signalling an otherwise largely unobserved impact of exposure to ambient air pollution. Understanding these patterns is essential to assess the full reproductive and population-level impact of environmental exposures and to inform maternal health policy and air quality regulation. Moreover, changes in the SRB may also have long-term demographic and societal implications. Skewed SRBs can affect future partnering dynamics, family formation patterns, and population dynamics, which may only become apparent decades later (Guilmoto, 2009). Potential changes in the SRB resulting from ambient air pollution exposure may therefore extend beyond reproductive health to public policy more broadly.

Ambient air pollution is already an urgent public health issue, and its burden is expected to worsen. Nearly the entire global population already lives in areas that do not meet WHO air quality guidelines (World Health Organization, 2021). While mechanisms are complex, climate change may intensify the air pollution, increasing ground-level ozone formation and fuelling wildfires, which contribute to elevated particulate matter (World Health Organization, 2021). Co-exposure to heat and air pollution may also have synergistic effects on adverse pregnancy outcomes (Raghavan *et al*., 2025). Rapid urbanisation, particularly in LMICs, is exposing growing populations to worsening air quality. These worsening air quality conditions may amplify any existing effects on SRB, making it imperative to understand these relationships now. This systematic review aimed to assess the available evidence on the relationship between ambient air pollution and the SRB.

## 2. Methods

The review protocol is registered on PROSPERO (no. CRD420250651445).

### 2.1 Search strategy details

We conducted a systematic review in accordance with the PRISMA guidelines (appendix pp.2-3). We searched MEDLINE, Embase, and Global Health (Ovid) on 12 February 2025, without language or date restrictions, using a combination of controlled vocabulary (MeSH for MEDLINE, Emtree for Embase) with free text search (by title and abstract for Global Health; by title, abstract and keywords for MEDLINE and Embase (see appendix p.4 for the full search). Air pollution search terms included (1). Criteria air pollutants, e.g., particulate matter (PM_2·5_, PM_10_, ultrafine particles), nitrogen dioxide and oxides, sulphur dioxide, ozone, carbon monoxide; (2) Hazardous Air Pollutants (e.g., Volatile organic compounds, polycyclic aromatic hydrocarbons, heavy metals persistent organic pollutants, and airborne pesticides or endocrine disrupting chemicals; (3) Emerging air pollutants (e.g., black and brown carbon, and microplastics). Air pollution exposure terms were combined with sex ratio search terms (e.g., “sex ratio”, “male-to-female birth ratio”, “proportion of male births”, “probability of male birth”).

### 2.2 Inclusion and exclusion criteria

We included original observational and quasi-experimental studies, including cohort studies (both prospective and retrospective), case-control studies, and cross-sectional designs, that assessed exposure to ambient air pollution and the SRB in human populations. We excluded studies that were not original research studies, such as conference abstracts, letters, editorials or reviews. Systematic and scoping reviews were not included in the final analysis but were used to check for additional eligible studies through reference screening.

Eligible studies were those that examined ambient air pollution as the primary exposure, specifically measuring concentrations of air pollutants (e.g., PM_2·5_, PM_10_), regardless of whether these were of anthropogenic or biogenic origin. Studies that measured other pollution sources (e.g., indoor or occupational exposures) to adjust for confounding were eligible, but studies that focused exclusively on these non-ambient sources, without including estimates of ambient air quality, were excluded. Eligible studies measured maternal exposure during a defined pre-conception window, during pregnancy, or aggregated exposure and birth outcomes by calendar year. We excluded studies that did not measure ambient pollution concentrations but discussed air pollution as a potential explanation for changes in the SRB. We also excluded studies that assessed toxin exposure via biomarkers (e.g., blood serum, bone lead levels) that could not be temporally linked to pregnancy or a pre-conception window, but that rather reflect long-term cumulative exposure over the life course.

Studies that reported pregnancy outcomes (miscarriage, stillbirth) without assessing the sex ratio were excluded, as were those focusing solely on the primary (conception) sex ratio and paternal exposures (e.g., assisted reproduction studies examining spermatogenesis). Finally, we excluded in vitro fertilisation (IVF) studies as these do not reflect in utero exposure.

### 2.3 Screening and data extraction

Two reviewers independently screened results by title and abstract followed by full text review. Where conflicts arose, decisions were adjudicated by a third reviewer. Rayyan was used to screen all search records. For eligible studies, we extracted the following information from each study: year of publication, study location, study design, sample size (women or births), gestational exposure window, gestational measurement, residence criteria, AQ assessment, average exposure in sample, outcome measure, type of particulate matter, spatial resolution of air quality data, temporal frequency of air quality data, air quality modelling, and effect size. Data were double extracted by two reviewers, and any discrepancies were resolved by a third reviewer.

### 2.4 Risk of bias

Study quality was appraised using an adapted version of the NIH Quality Assessment Tool for Observational Cohort and Cross-Sectional Studies (NHLBI 2014), modified to accommodate ecological, quasi-experimental, and time-series designs. The adapted criteria evaluated study design, exposure and outcome assessment quality, control for confounding, statistical methods, and risk of bias (see appendix p.4). Two reviewers completed the quality appraisal, with discrepancies resolved by a third reviewer.

### 2.5 Data analysis

We used a narrative synthesis to summarise findings and an effect direction plot to show whether each study shows masculinisation, feminisation, or no change in the SRB with exposure to ambient pollution. This enabled a consistent presentation of effect direction across studies employing different outcome and effect measures. Meta-analysis was infeasible due to the limited number of eligible studies, combined with considerable heterogeneity in study design, pollutant types, exposure categorisations, pre-conception and pregnancy exposure windows, and spatial attribution (Valentine, Pigott and Rothstein, 2010).

## 3. Results

### 3.1 Study selection

The study selection process is illustrated in **Figure 2** (PRISMA diagram). A total of 592 records were identified through database searches (Embase: 328; MEDLINE: 232; Global Health: 32). After removing duplicates, 421 unique records remained. Title and abstract screening reduced this to 52 studies selected for full-text review, of which 11 met the inclusion criteria.

**Figure 2.**
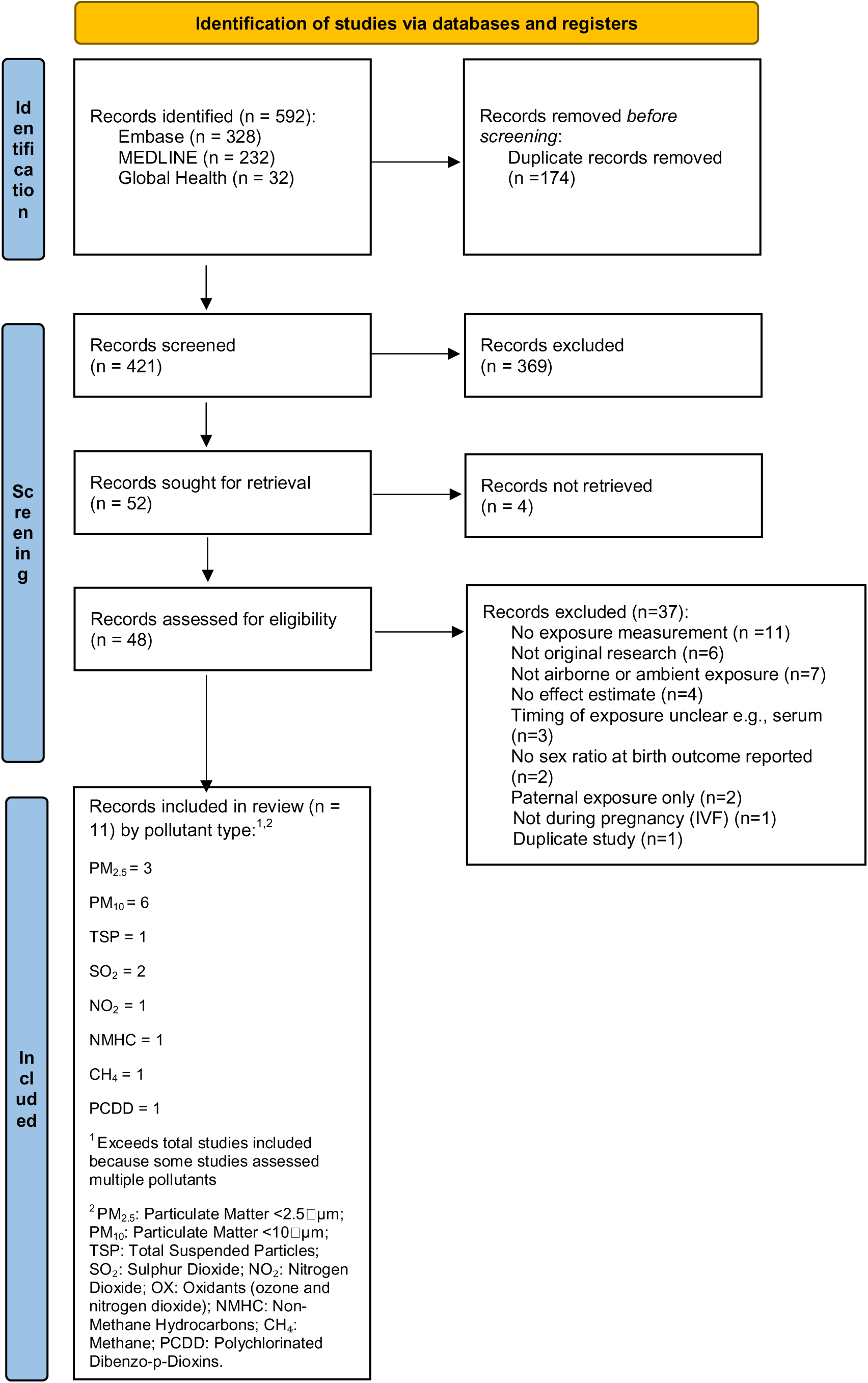
PRISMA flow diagram.

### 3.2 Study characteristics

Table 1 presents the characteristics of the included studies. Of the 11 included studies, the years of observation spanned from 1970 to 2023, although most studies were published within the last decade. Sample sizes varied substantially, ranging from small cohorts to national-level datasets exceeding one million births (e.g., Great Britain (Ghosh *et al*., 2019, p. 20), USA (Sanders and Stoecker, 2015)). Several studies examined air quality for a population defined by their proximity to specific sources of ambient pollution, such as municipal waste incinerators (Lin, Li and Mao, 2006; Candela *et al*., 2013; Santoro *et al*., 2016; Ghosh *et al*., 2019). The evidence base was geographically narrow, with studies conducted in only eight countries, all of which were either high-income or upper-middle-income economies. Brazil and China were the only middle-income countries represented. Entire regions, including South Asia and Sub-Saharan Africa, were not represented in any of the existing evidence.

**Table 1.** Study characteristics.

| Study | Study location | Sample detail | Observation period | Sample size births | Type of study | Pollutant source | Pollutants assessed |
| --- | --- | --- | --- | --- | --- | --- | --- |
| Arima et al (2023) <sup>1</sup> | Fukue Island, Japan | Not reported | Pollutants: 2013-2022<br>Births: 2014- 2022 | 1,835 | Ecological (population-level, time-series) | Ambient | PM<sub>2.5</sub>, SO<sub>2</sub>, OX |
| Candela et al (2013) <sup>2</sup> | Emilia-Romagna, Italy | 4km radius around eight incinerators | 2003-2010 | 21,164 | Observational (individual-level, retrospective cohort) | Municipal waste incinerators (MWI) | PM<sub>10</sub> |
| Ghosh et al (2019) <sup>3</sup> | Great Britain | 22 municipal waste incinerators | 2003-2010 | 1,025,064 | Matched case control (individual-level) | MWI | PM<sub>10</sub> |
| Goin et al (2024) <sup>4</sup> | Paradise, California, USA | Not reported | 2017-2019 | Not reported | Quasi-experimental (individual-level, natural experiment) | Wildfire | PM<sub>2.5</sub> |
| Houdek et al (2020) <sup>5</sup> | Czech Republic | Residents of 216 districts | 1992-2010 | 446,411 | Observational (individual-level, retrospective cohort) | Ambient | PM<sub>10</sub> |
| Lin et al (2006) <sup>6</sup> | Taipei, Taiwan | One MWI | 1991, 1997 | 1991: 6,697<br>1997: 6,282 | Observational (ecological exposure, individual-level covariates, cross-sectional pre-post comparison) | MWI | PCDDs |
| Lin et al (2015) <sup>7</sup> | Guangzhou, China | Births in one major maternity hospital | 2006-2011 | 81,711 | Observational: (individual-level, retrospective cohort) | Ambient | PM<sub>10</sub>, NO<sub>2</sub>, SO<sub>2</sub> |
| Miraglia et al (2013) <sup>8</sup> | Sao Paulo, Brazil | - | 2000-2007 | 53,612 | Ecological (population-level, time-series) | Ambient | PM<sub>10</sub> |
| Sanders & Stoecker (2015) <sup>9</sup> | USA | 198 counties, 35 states | 1970-1973 | 2,709,415 | Quasi-experimental (population-level, natural experiment) | Ambient | TSPs |
| Santoro et al (2016) <sup>10</sup> | Arezzo, Tuscany, Italy | One incinerator plant | 2001-2010 | 3,069 | Observational (individual-level, retrospective cohort) | MWI | PM<sub>10</sub> |
| Tsai et al (2025) <sup>11</sup> | Taipei, Taiwan | Not reported | 1992-2023 | Period 1: 137,438<br>Period 2: 263,016<br>Period 3: 261,708<br>Period 4: 163,630 | Ecological (population-level, time series) | Ambient, pre-post implementation of a rapid transit system | PM<sub>2.5</sub> |

Across studies, PM□□ was the most frequently studied air pollutant (six studies), followed by PM_2·5_ (three studies). All other air pollutants were examined in only one or two studies.

Study designs used to investigate the relationship between SRB and air quality were highly heterogeneous. Three studies used ecological, population-level approaches (Miraglia *et al*., 2013; Arima, 2023; Tsai, Weng and Yang, 2025). Two adopted quasi-experimental designs, with wildfire (Goin *et al*., 2024) or regulatory change used as an exogenous shock to air quality (Sanders and Stoecker, 2015).

### 3.3 Exposure and outcome measures

Table 2 summarises how air pollution exposures and the sex ratio at birth were measured. There was considerable variation in the temporal resolution of exposure assessment. Seven studies relied on aggregate-level data (Lin, Li and Mao, 2006; Miraglia *et al*., 2013; Sanders and Stoecker, 2015; Santoro *et al*., 2016; Houdek, Dvouletý and Pažitka, 2020; Arima, 2023; Tsai, Weng and Yang, 2025), typically assigning exposure based on annual births without aligning exposure windows to the gestational period. Two studies tracked exposure throughout pregnancy but summarised this into a cumulative average of exposure across the woman’s entire pregnancy (Candela *et al*., 2013; Ghosh *et al*., 2019), precluding analysis of critical windows of gestational exposure. Two studies assigned exposure with greater temporal specificity: one assessed daily exposure in the two weeks prior to conception (Lin *et al*., 2015), while another examined the timing of wildfire events relative to gestational age to identify critical windows of vulnerability (Goin *et al*., 2024).

**Table 2.** Exposure and outcome measures.

| **Study** | **Exposure window**<br>**(in relation to**<br>**pregnancy)** | **Frequency of**<br>**exposure**<br>**measurement**<br>**throughout**<br>**pregnancy** | **Spatial inclusion**<br>**criteria** | **Exposure**<br>**attribution** | **AQ definition** | **Available**<br>**temporal**<br>**frequency of AQ**<br>**data** | **Sex ratio outcome**<br>**measure** |
| --- | --- | --- | --- | --- | --- | --- | --- |
| **Arima et al (2023)<sup>1</sup>** | Year of birth | NA (gestational window not measured) | Not specified | Area-level | Monthly mean exposure during daylight hours | Hourly aggregated to monthly | Male-to-female ratio at birth |
| **Candela et al (2013)<sup>2</sup>** | Entire pregnancy | Summary average over pregnancy | Mothers' residence within 4km radius around MWI | Individual-level | Categorical pregnancy-average, quintiles | Half-hourly (~17,000 measures/year) | Proportion female births (female / total) |
| **Ghosh et al (2019)<sup>3</sup>** | Entire pregnancy | Summary average over pregnancy | Mothers' residence within 10km radius around MWI | Individual-level | Mean daily exposure across pregnancy | Daily aggregated to pregnancy average | Female-to-male ratio at birth |
| **Goin et al (2024)<sup>4</sup>** | Dates of wildfire during pregnancy | Weekly | Mothers' residence within census block group centroid and plume polygons | Individual-level | Binary exposure (thresholds exceeded and duration) | Hourly aggregated to daily | Male-to-female ratio at birth |
| **Houdek et al (2020)<sup>5</sup>** | Month of birth | Summary average over month of birth | District-level (216 stations) | Area-level | Monthly average | Daily aggregated to monthly | Binary outcome: male birth |
| **Lin et al (2006)<sup>6</sup>** | Year of birth | NA (gestational window not measured) | Not specified | Area-level | Categorical annual exposure (reference, medium, high) | Hourly aggregated to annual | Binary outcome: female birth |
| **Lin et al (2015)<sup>7</sup>** | 13 days pre-conception | Daily | City-wide average (9 monitors) | Area-level | Daily average | Daily | Binary outcome: female birth |
| **Miraglia et al (2013)<sup>8</sup>** | Year of birth | NA (gestational window not measured) | City-wide average (9 monitors) | Area-level | Annual average | Daily aggregated to annual | Proportion male births (male / total) |
| **Sanders & Stoecker (2015)<sup>9</sup>** | Year of birth | NA (gestational window not measured) | County-level | Area-level | Annual exposure (attainment status) | Daily aggregated to annual | Proportion male births (male / total) |
| **Santoro et al (2016)<sup>10</sup>** | Year of birth | NA (gestational window not measured) | Mothers' residence within 12km around MWI | Individual-level | Categorical annual exposure (low/medium/high) | Annual | Male-to-female birth ratio |
| **Tsai et al (2025)<sup>11</sup>** | Year of birth | NA (gestational window not | City-wide average (6 monitors) | Area-level | Annual average PM2.5 (estimated | Hourly data aggregated to daily | Proportion male births (male / total) |
|  |  | measured) |  |  | from PM10) | and annual |  |

Only four studies assigned exposure at the individual level (Candela *et al*., 2013; Santoro *et al*., 2016; Ghosh *et al*., 2019; Goin *et al*., 2024), typically using the mother’s residential postcode at birth and birth date to attribute air pollution levels during the pregnancy. The remaining studies attributed exposure at the area level, applying average pollution estimates across entire cities, districts, or counties (Lin, Li and Mao, 2006; Miraglia *et al*., 2013; Lin *et al*., 2015; Sanders and Stoecker, 2015; Houdek, Dvouletý and Pažitka, 2020; Arima, 2023; Tsai, Weng and Yang, 2025). Exposure was treated either as a continuous variable (for example, daily, monthly, or annual mean pollution levels) (Miraglia *et al*., 2013; Lin *et al*., 2015; Sanders and Stoecker, 2015; Ghosh *et al*., 2019; Houdek, Dvouletý and Pažitka, 2020; Arima, 2023; Tsai, Weng and Yang, 2025), categorical (e.g., quintiles) (Lin, Li and Mao, 2006; Candela *et al*., 2013; Santoro *et al*., 2016), or binary thresholds exceedances to distinguish exposed from unexposed areas (Goin *et al*., 2024).

### 3.4 Main findings

The main findings are presented in **Table 3** (by study) and **Figure 3** (by air pollutant type).

**Figure 3.**
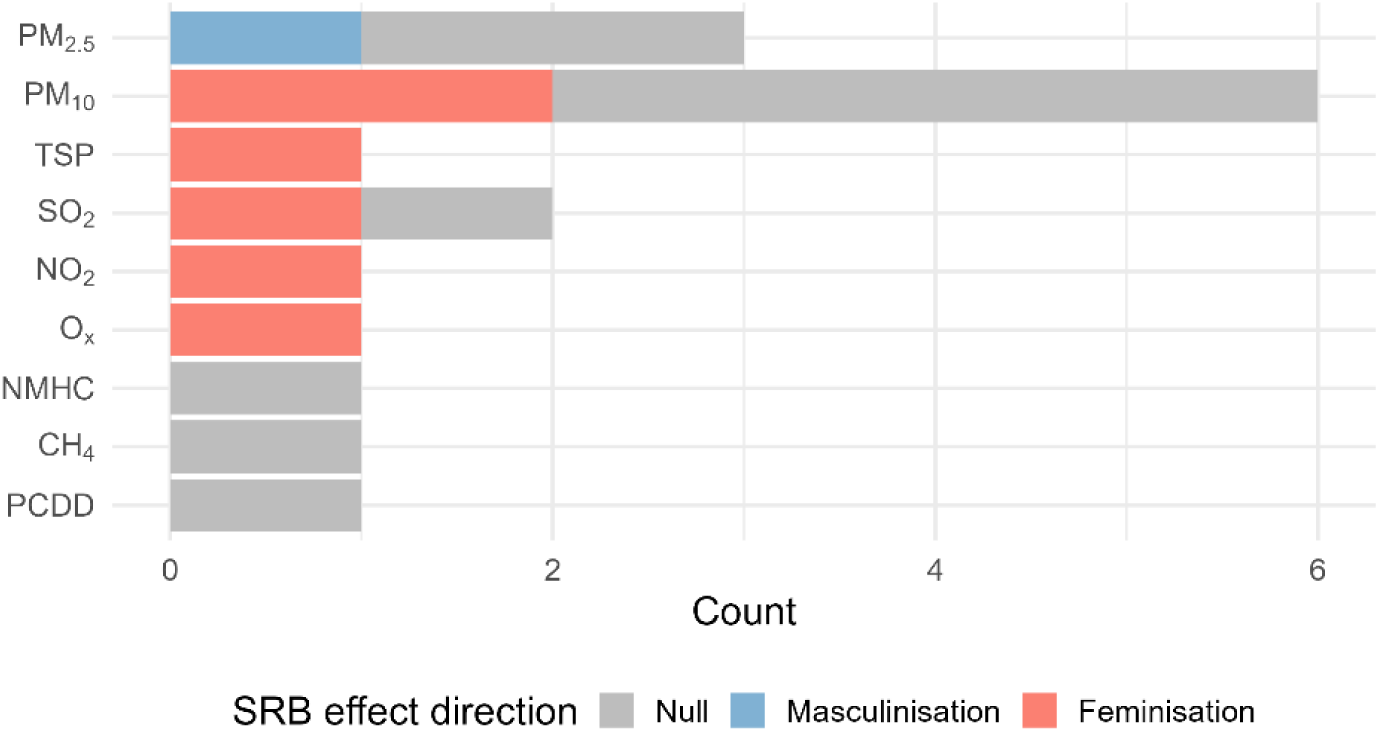
Effect direction counts.

**Table 3.**
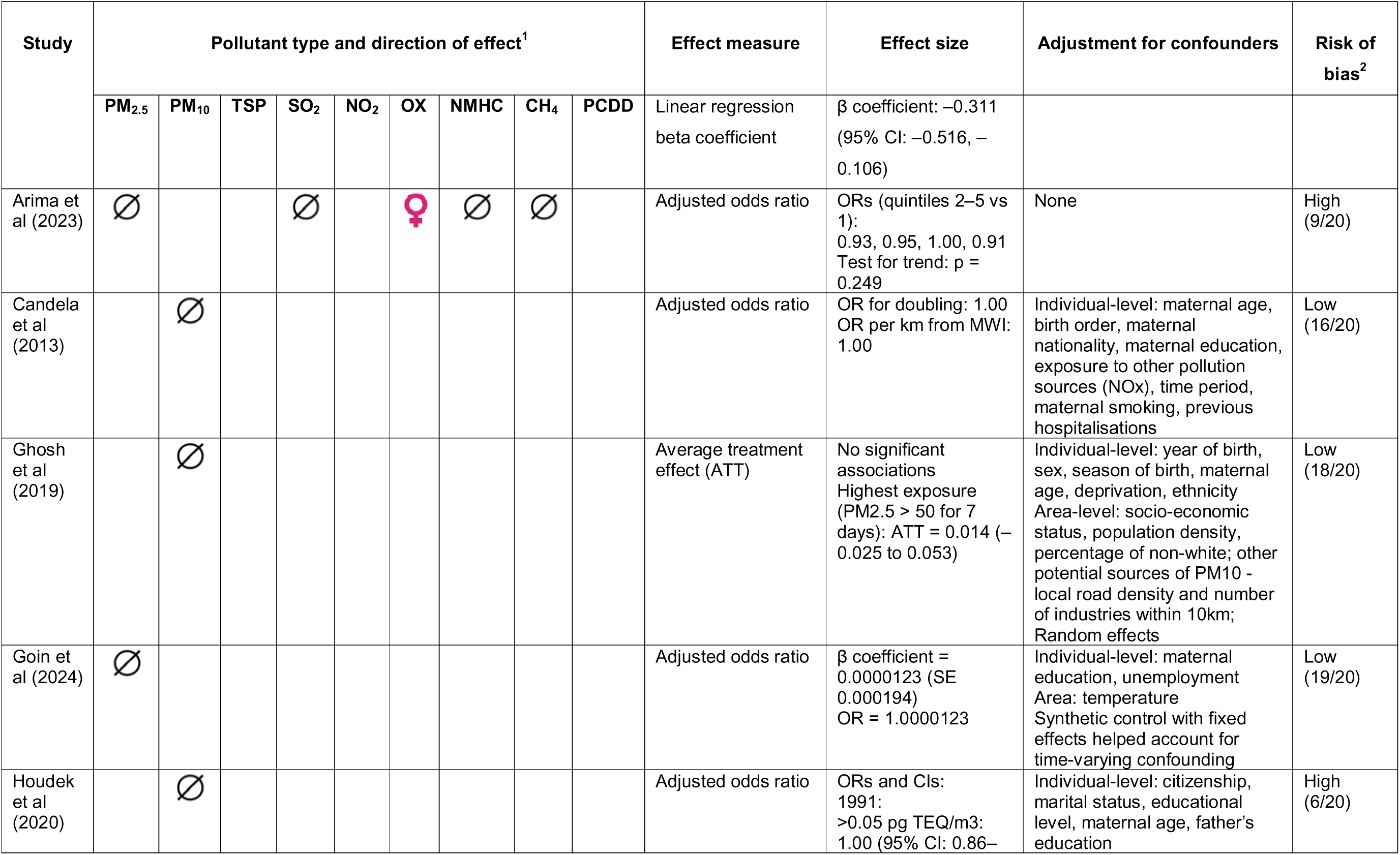
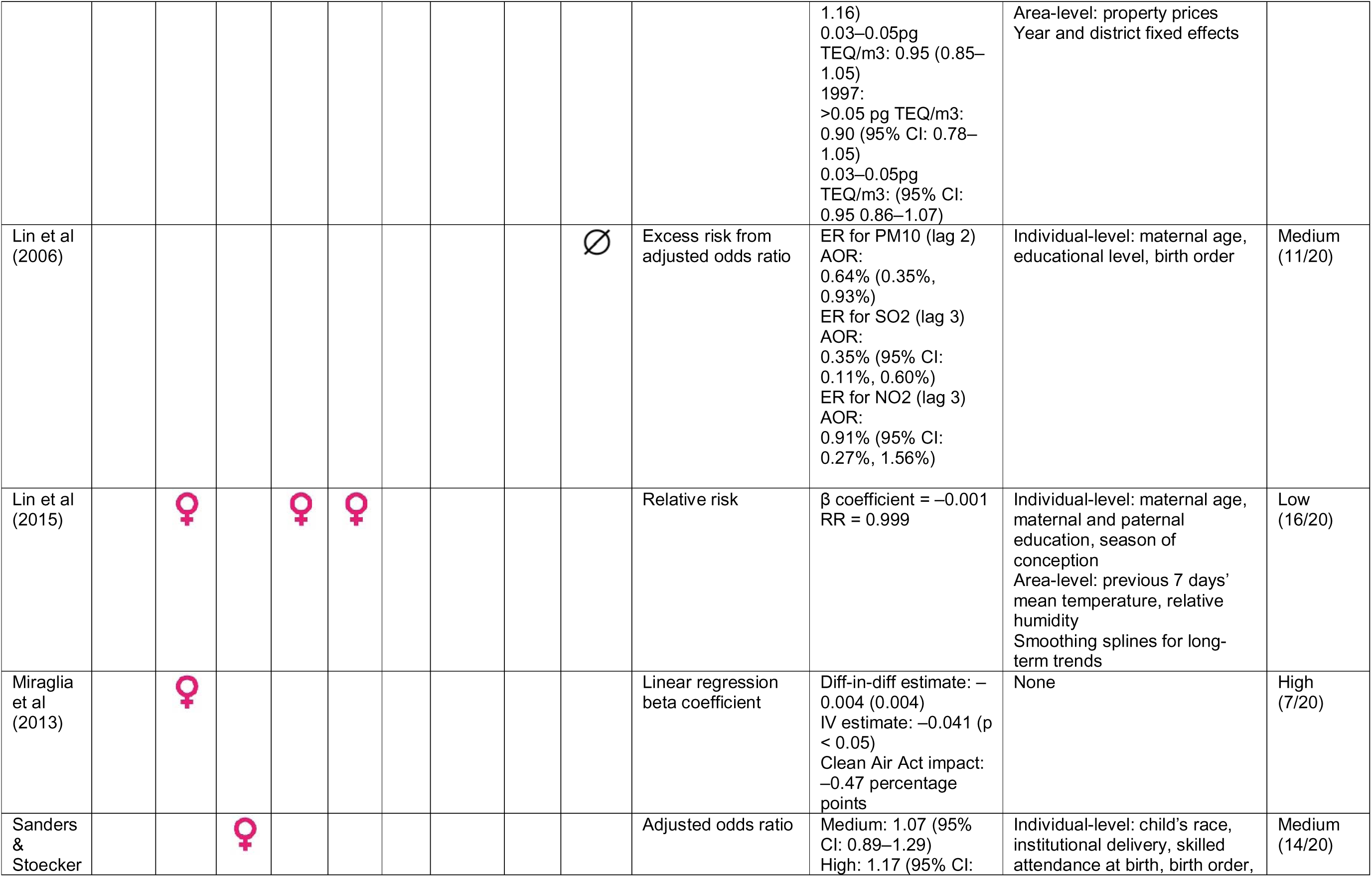
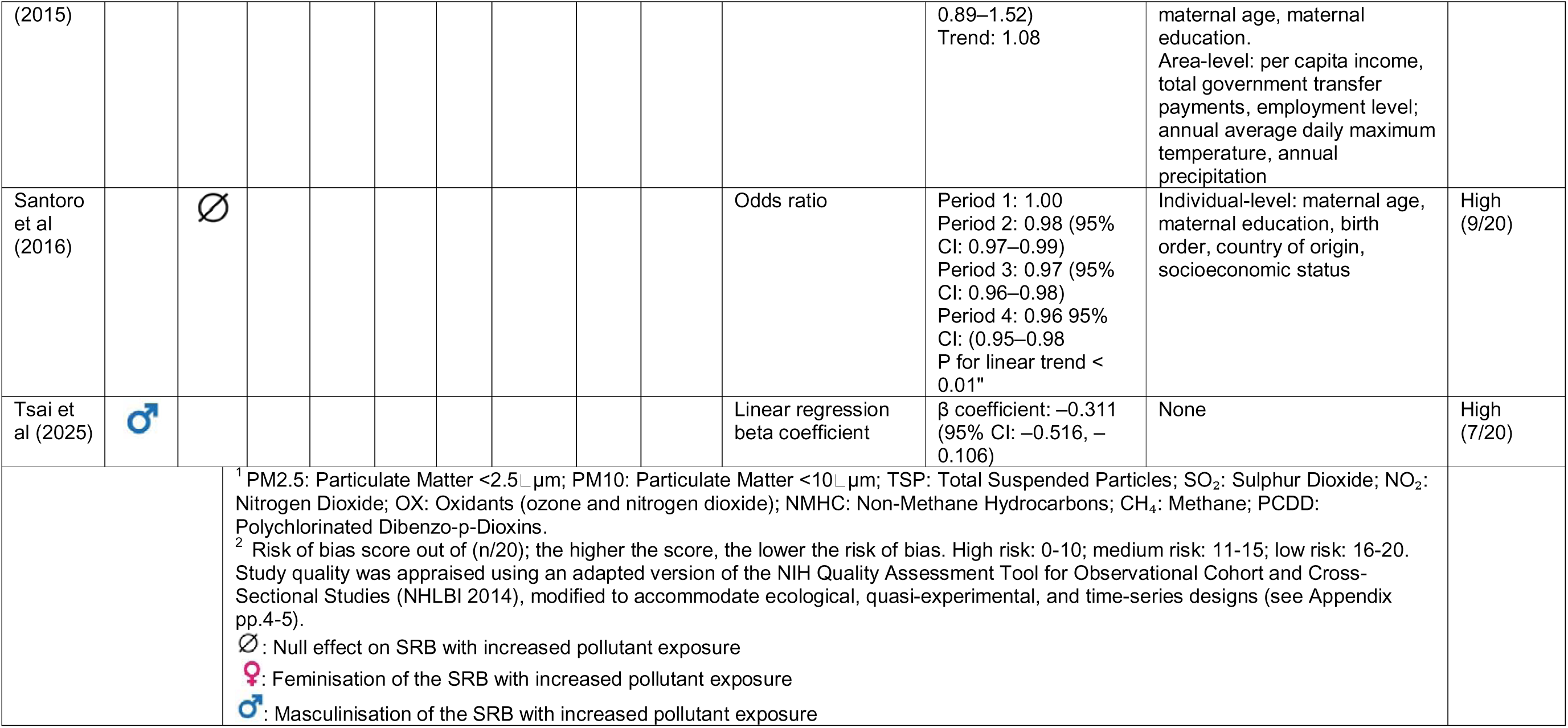
Main results and risk of bias.

Four studies reported that higher levels of air pollution were associated with a feminisation of the SRB (Miraglia *et al*., 2013; Lin *et al*., 2015; Sanders and Stoecker, 2015; Arima, 2023). This association was observed for various pollutants, including PM_10_ (Miraglia *et al*., 2013; Lin *et al*., 2015), total suspended particulates (TSP) (Sanders and Stoecker, 2015), sulphur dioxide (SO□) (Lin *et al*., 2015), nitrogen dioxide (NO□) (Lin *et al*., 2015), and oxides (Arima, 2023). One study found that increased PM_2·5_ exposure was associated with a masculinisation of the SRB (Tsai, Weng and Yang, 2025).The remaining six studies reported only null findings, either for one or multiple pollutants (Lin, Li and Mao, 2006; Candela *et al*., 2013; Santoro *et al*., 2016; Ghosh *et al*., 2019; Houdek, Dvouletý and Pažitka, 2020; Goin *et al*., 2024, p. 202).

Eight studies adjusted for some individual-level confounders (Lin, Li and Mao, 2006; Candela *et al*., 2013; Lin *et al*., 2015; Sanders and Stoecker, 2015; Santoro *et al*., 2016; Ghosh *et al*., 2019; Houdek, Dvouletý and Pažitka, 2020; Goin *et al*., 2024), five of which also included area-level covariates. Only three studies included other climatic stressors (Lin *et al*., 2015; Sanders and Stoecker, 2015; Goin *et al*., 2024), such as temperature and humidity, which influence pollutant dispersion and intensity, as well as pregnancy outcomes. However, only one study accounted for maternal health status, including smoking patterns and maternal morbidity (Candela *et al*., 2013). Potential confounding variables such as maternal smoking and body mass index (BMI) were not adjusted for in the other ten studies, even though they may be associated with socioeconomic status, air pollution exposure, and potentially sex-biased pregnancy loss. Similarly, although two studies assessed industrial pollution alongside ambient exposures (Candela *et al*., 2013; Ghosh *et al*., 2019), none examined potential confounding from other sources such as indoor cooking fuels or occupational exposures.

### 3.5 Risk of bias

Risk of bias scores are presented in Table 3. On our modified 20-point scale, five studies were classified as high risk of bias (≤10) (Miraglia *et al*., 2013; Santoro *et al*., 2016; Houdek, Dvouletý and Pažitka, 2020; Arima, 2023; Tsai, Weng and Yang, 2025), two as medium risk (11–15) (Lin, Li and Mao, 2006; Sanders and Stoecker, 2015), and four as low risk (16–20) (Candela *et al*., 2013; Lin *et al*., 2015, p. 20; Ghosh *et al*., 2019; Goin *et al*., 2024) (see appendix p.5 for score breakdown). The most common limitations were related to exposure misclassification, particularly in ecological studies that aggregated births within a given period (most commonly by calendar year) and measured air quality over the same period, without attributing exposure to the gestational window at the individual level. Additional concerns included weak causal inference due to study design limitations, and restricted sample representativeness, for instance when geographically restricted to women residing within a small radius of an MWI (Candela *et al*., 2013; Santoro *et al*., 2016; Ghosh *et al*., 2019).

### 3.6 Biological mechanisms

Across the 11 studies, the mechanisms through which exposure to ambient air pollution may affect the SRB were largely unexplored. Only one study, Lin et al (2015) explicitly examined maternal exposure in a defined pre-conception window, up to 13 days before the estimated conception date (Lin *et al*., 2015). Three studies assessed maternal exposure during pregnancy at the individual-level and inferred sex-differentials in early conceptus and pregnancy loss based on shifts in the SRB. However, two of these aggregated maternal exposure into a single summary measure, precluding the identification of windows of gestational vulnerability or the effect of chronic versus acute exposure (Candela *et al*., 2013; Ghosh *et al*., 2019). In the remaining seven studies, birth outcomes were aggregated over calendar periods without temporally aligning exposure to specific gestational windows, also limiting the ability to identify mechanisms underlying changes in the SRB (Lin, Li and Mao, 2006; Miraglia *et al*., 2013; Sanders and Stoecker, 2015; Santoro *et al*., 2016; Houdek, Dvouletý and Pažitka, 2020; Arima, 2023; Tsai, Weng and Yang, 2025).

## 4. Discussion

The SRB is a sentinel marker of reproductive stress at the population level, reflecting the cumulative impact of sex-biased conception and pregnancy losses that would otherwise remain largely unobserved. Despite growing public health concern about ambient air pollution, and the large proportion of the world’s population already exposed to air pollution that exceeds WHO standards (World Health Organization, 2021), its association with the SRB remains underexplored and poorly understood. To our knowledge, this is the first systematic review focused specifically on the contribution of air pollution to the SRB.

From the 11 studies included in our review, the evidence for the effect of air pollution on the SRB is limited, highly heterogeneous, and largely inconclusive. This finding is consistent with an existing review of environmental pollutants (Azizi *et al*., 2024). Most studies reported null findings, including those with the lowest risk of bias (Ghosh *et al*., 2019; Goin *et al*., 2024). Four studies found weak evidence of a feminisation of the SRB associated with exposure to ambient air pollution (Miraglia *et al*., 2013; Lin *et al*., 2015; Sanders and Stoecker, 2015; Arima, 2023), though these studies varied in their risk of bias. Feminisation of live births, resulting in a reduced SRB, is consistent with the hypothesis that male conceptuses are more likely to be spontaneously terminated under maternal stress (Bruckner and Catalano, 2018; Catalano, Casey and Bruckner, 2020). Evidence that ambient air pollution differentially increases male conceptus loss also supports the Trivers-Willard hypothesis, which predicts that adverse conditions favour the survival of female offspring (Trivers and Willard, 1973). However, heterogeneity in pollutant types, exposure windows, effect measures, and study design precluded meta-analysis to estimate the direction and strength of the association between air pollution and the SRB. There is an urgent need for new primary studies using standardised methods to improve comparability and facilitate future meta-analyses.

The existing evidence is geographically narrow, limited to only eight countries (Italy, China, Taiwan, Czech Republic, Brazil, the UK, Japan, and the USA) and lacks data from regions with some of the world’s poorest air quality and greatest burden of ambient air pollution attributable morbidity and mortality, including South Asia and sub-Saharan Africa.(WHO, 2025) Existing research also focuses mainly on particulate matter (PM_10_ and PM_2·5_), with minimal examination of other pollutants such as ozone (O□), nitrogen oxides (NO□), or sulphur dioxide (SO□). This is a critical gap, as co-exposure to multiple pollutants may influence toxicity and reproductive health in complex ways (Chen *et al*., 2024; He *et al*., 2025). For instance, PM_2·5_ and O□ co-exposure may synergistic effects exceeding either pollutant alone (He *et al*., 2025), yet such interactions remain largely unexplored for pregnancy loss, including as indicated at the population-level by shifts in the SRB. Moreover, it is unclear which levels of air pollution are harmful for the SRB or whether associations follow a linear dose–response--information that is essential for policy design.

Methodological quality across the 11 studies was highly variable, five studies rated as having a high risk of bias. Many were limited by exposure timing, exposure misclassification, and omission of confounders. Seven studies used aggregate birth outcome data (Lin, Li and Mao, 2006; Miraglia *et al*., 2013; Sanders and Stoecker, 2015; Santoro *et al*., 2016; Houdek, Dvouletý and Pažitka, 2020; Arima, 2023; Tsai, Weng and Yang, 2025) (e.g., by year), without linking exposure to individual pregnancies, precluding temporal alignment and identification of critical windows of vulnerability. Among the four studies with exposure aligned with pregnancy timing, two still used cumulative averages across the entire gestation (Candela *et al*., 2013; Ghosh *et al*., 2019). Only two studies examined exposure with greater temporal specificity: one assessed daily exposure in the two weeks prior to conception (Lin *et al*., 2015), and another examined wildfire events relative to gestational age (Goin *et al*., 2024).

Potential exposure misclassification was also common. Seven studies relied on area-level exposure from fixed-site monitors, with accuracy depending on area size and spatial variability of pollutant concentrations (Lin, Li and Mao, 2006; Miraglia *et al*., 2013; Lin *et al*., 2015; Sanders and Stoecker, 2015; Houdek, Dvouletý and Pažitka, 2020; Arima, 2023; Tsai, Weng and Yang, 2025). Studies that estimated individual-level attribution with greater spatial specificity used air quality models that incorporated maternal residence and pollutant dispersion influenced by emission sources, distance, climate and topography (Candela *et al*., 2013; Santoro *et al*., 2016; Ghosh *et al*., 2019; Goin *et al*., 2024). However, these assumed a fixed residence throughout pregnancy and did not capture exposure occurring away from home. Misclassification may persist, particularly in high pollution settings where pregnant individuals modify mobility patterns to avoid harmful air.

Third, the omission of potential confounders was common. Key maternal health characteristics, including smoking and maternal morbidities, were omitted from all but one study (Candela *et al*., 2013). No study included body mass index (BMI). Maternal smoking (Parazzini *et al*., 2005; Beratis, Asimacopoulou and Varvarigou, 2008; Koshy *et al*., 2010) and low or high maternal BMI (Cagnacci *et al*., 2004; DeVilbiss *et al*., 2022) may correlate with deprivation and residence in more polluted areas as well as pregnancy loss. Only two studies controlled for seasonality (Lin *et al*., 2015; Ghosh *et al*., 2019), despite strong seasonal variation in both air pollution and births. Few studies addressed spatial confounding – only one used random effects (Ghosh *et al*., 2019, p. 201), another applied district fixed effects (Houdek, Dvouletý and Pažitka, 2020), and one employed a generalised synthetic control approach to address both time-varying and invariant confounding (Goin *et al*., 2024).

Finally, the role of heat in the association between air pollution and the SRB warrants further attention. With a few exceptions (Lin *et al*., 2015; Sanders and Stoecker, 2015; Goin *et al*., 2024), most studies did not consider temperature or explore how it should be treated in statistical models. Depending on causal direction, temperature could act as a confounder, a mediator, or an effect modifier. Interest in how heat and air pollution interact to affect health outcomes is growing, yet this issue remains largely unexamined in relation to the SRB.

### 4.1 Biological mechanisms

Across the 11 studies, biological mechanisms were underexplored and remain difficult to disentangle. Observed shifts in the SRB may reflect (1) sex-biased conception, influenced by maternal and/or paternal factors such as oocyte or sperm quality, or (2) sex-biased pregnancy loss, encompassing implantation failures as well as differential survival of male versus female conceptuses throughout gestation. The first pathway is challenging to investigate outside of assisted reproduction settings. Most population-based studies, which estimate maternal exposures retrospectively from live births, are better positioned to capture pregnancy loss mechanisms. Only one study explicitly examined pre-conception mechanisms (Lin *et al*., 2015), which may reflect a effects on sperm quality through oxidative DNA damage or epigenetic changes (James, 2001; Van Larebeke *et al*., 2008; Radwan *et al*., 2018). Three studies investigated post-conception exposures consistent with pregnancy loss mechanisms but did not identify specific biological pathways (Candela *et al*., 2013, p. 20; Ghosh *et al*., 2019; Goin *et al*., 2024). Air pollutants can trigger oxidative stress, resulting in DNA damage to gametes or embryonic tissues (Carré *et al*., 2017), they may interfere with endocrine signalling that is critical for sex-specific embryonic development; and they can impair placental function, which differs by foetal sex and may render male foetuses more susceptible to adverse intrauterine conditions (Clifton, 2010; Braun *et al*., 2022). For the remaining seven studies, they did not attempt to disentangle the contribution of conception effects and differential pregnancy loss on the SRB (Lin, Li and Mao, 2006; Miraglia *et al*., 2013; Sanders and Stoecker, 2015; Santoro *et al*., 2016; Houdek, Dvouletý and Pažitka, 2020; Arima, 2023; Tsai, Weng and Yang, 2025). Advancing understanding of these pathways is essential to interpret observed SRB changes and identify when, how, and in whom ambient air pollution affects reproductive outcomes.

### 4.2 Strengths and limitations

This systematic review has several strengths, including a comprehensive search strategy, double screening and extraction, and a conceptual framework linking ambient air pollution to the SRB. However, we could not conduct a meta-analysis or assess publication bias due to the limited, heterogeneous evidence base, underscoring the need for additional primary research. Future studies should prioritise: (1) identifying critical windows of exposure; (2) using individual-level data with accurate exposure measures and key confounders (e.g. seasonality and place specific characteristics); (3) expanding studies to high-pollution, underrepresented regions, especially sub-Saharan Africa and South Asia; (4) incorporating pollutant combinations and investigating the role of heat as an effect modifier, modifier or confounder; and (5) directly investigating biological mechanisms through biomarkers of oxidative stress, placental function, and endocrine disruption.

As ambient air quality deteriorates in a rapidly warming planet, understanding how air pollution affects the SRB is increasingly important. As a sentinel indicator of reproductive health, the SRB captures the otherwise unobserved, cumulative impact of sex-biased conception and pregnancy loss. Strengthening the evidence on this association is critical to inform air quality regulation and guide public health responses that protect reproductive health.

## Data Availability

All studies included in the systematic review are available in the public domain

## Acknowledgements

None.

## Declarations

Data Sharing

N/A all studies are available in the public domain.

Declaration of interests

We declare no competing interests.

Declaration of generative AI and AI-assisted technologies in the manuscript preparation process

During the preparation of this work the authors used ChatGPT to assist with grammar and language editing. After using this tool/service, the authors reviewed and edited the content as needed and take full responsibility for the content of the published article.

