## Supplemental material for "Ambient air pollution and the sex ratio at birth: a systematic review and narrative synthesis"

**Table S1. PRISMA checklist**

| **Section and Topic** | **Item #** | **Checklist item** | **Location where item is reported** |
| --- | --- | --- | --- |
| **TITLE** |  |  |  |
| Title | 1 | Identify the report as a systematic review. | p.1 |
| **ABSTRACT** |  |  |  |
| Abstract | 2 | See the PRISMA 2020 for Abstracts checklist. | p.2 |
| **INTRODUCTION** |  |  |  |
| Rationale | 3 | Describe the rationale for the review in the context of existing knowledge. | pp.5-6 |
| Objectives | 4 | Provide an explicit statement of the objective(s) or question(s) the review addresses. | p.6 |
| **METHODS** |  |  |  |
| Eligibility criteria | 5 | Specify the inclusion and exclusion criteria for the review and how studies were grouped for the syntheses. | pp.8-9 |
| Information sources | 6 | Specify all databases, registers, websites, organisations, reference lists and other sources searched or consulted to identify studies. Specify the date when each source was last searched or consulted. | p.8 |
| Search strategy | 7 | Present the full search strategies for all databases, registers and websites, including any filters and limits used. | Appendix |
| Selection process | 8 | Specify the methods used to decide whether a study met the inclusion criteria of the review, including how many reviewers screened each record and each report retrieved, whether they worked independently, and if applicable, details of automation tools used in the process. | p.9 |
| Data collection process | 9 | Specify the methods used to collect data from reports, including how many reviewers collected data from each report, whether they worked independently, any processes for obtaining or confirming data from study investigators, and if applicable, details of automation tools used in the process. | p.9 |
| Data items | 10a | List and define all outcomes for which data were sought. Specify whether all results that were compatible with each outcome domain in each study were sought (e.g. for all measures, time points, analyses), and if not, the methods used to decide which results to collect. | p.9 |
|  | 10b | List and define all other variables for which data were sought (e.g. participant and intervention characteristics, funding sources). Describe any assumptions made about any missing or unclear information. | p.9 |
| Study risk of bias assessment | 11 | Specify the methods used to assess risk of bias in the included studies, including details of the tool(s) used, how many reviewers assessed each study and whether they worked independently, and if applicable, details of automation tools used in the process. | p.9 |
| Effect measures | 12 | Specify for each outcome the effect measure(s) (e.g. risk ratio, mean difference) used in the synthesis or presentation of results. | p.9 |
| Synthesis methods | 13a | Describe the processes used to decide which studies were eligible for each synthesis (e.g. tabulating the study intervention characteristics and comparing against the planned groups for each synthesis (item #5)). | N/A |
|  | 13b | Describe any methods required to prepare the data for presentation or synthesis, such as handling of missing summary statistics, or data conversions. | N/A |
|  | 13c | Describe any methods used to tabulate or visually display results of individual studies and syntheses. | p.9 |
|  | 13d | Describe any methods used to synthesize results and provide a rationale for the choice(s). If meta-analysis was performed, describe the model(s), method(s) to identify the presence and extent of statistical heterogeneity, and software package(s) used. | p.9 |
|  | 13e | Describe any methods used to explore possible causes of heterogeneity among study results (e.g. subgroup analysis, meta-regression). | N/A |
|  | 13f | Describe any sensitivity analyses conducted to assess robustness of the synthesized results. | N/A |
| Reporting bias assessment | 14 | Describe any methods used to assess risk of bias due to missing results in a synthesis (arising from reporting biases). | p.9 |
| Certainty assessment | 15 | Describe any methods used to assess certainty (or confidence) in the body of evidence for an outcome. | N/A |
| **RESULTS** |  |  |  |
| Study selection | 16a | Describe the results of the search and selection process, from the number of records identified in the search to the number of studies included in the review, ideally using a flow diagram. | pp.10-11 |
|  | 16b | Cite studies that might appear to meet the inclusion criteria, but which were excluded, and explain why they were excluded. | p.11 |
| Study characteristics | 17 | Cite each included study and present its characteristics. | p.13 |
| Risk of bias in studies | 18 | Present assessments of risk of bias for each included study. | p.18 |
| Results of individual studies | 19 | For all outcomes, present, for each study: (a) summary statistics for each group (where appropriate) and (b) an effect estimate and its precision (e.g. confidence/credible interval), ideally using structured tables or plots. | p.15 |
| Results of syntheses | 20a | For each synthesis, briefly summarise the characteristics and risk of bias among contributing studies. | p.18 |
|  | 20b | Present results of all statistical syntheses conducted. If meta-analysis was done, present for each the summary estimate and its precision (e.g. confidence/credible interval) and measures of statistical heterogeneity. If comparing groups, describe the direction of the effect. | p.19 |
|  | 20c | Present results of all investigations of possible causes of heterogeneity among study results. | N/A |
|  | 20d | Present results of all sensitivity analyses conducted to assess the robustness of the synthesized results. | N/A |
| Reporting biases | 21 | Present assessments of risk of bias due to missing results (arising from reporting biases) for each synthesis assessed. | N/A |
| Certainty of evidence | 22 | Present assessments of certainty (or confidence) in the body of evidence for each outcome assessed. | N/A |
| **DISCUSSION** |  |  |  |
| Discussion | 23a | Provide a general interpretation of the results in the context of other evidence. | p.20 |
|  | 23b | Discuss any limitations of the evidence included in the review. | pp.20-22 |
|  | 23c | Discuss any limitations of the review processes used. | p.22 |
|  | 23d | Discuss implications of the results for practice, policy, and future research. | pp.20, 22 |
| **OTHER INFORMATION** |  |  |  |
| Registration and protocol | 24a | Provide registration information for the review, including register name and registration number, or state that the review was not registered. | pp.2,8 |
|  | 24b | Indicate where the review protocol can be accessed, or state that a protocol was not prepared. | pp.2,8 |
|  | 24c | Describe and explain any amendments to information provided at registration or in the protocol. | N/A |
| Support | 25 | Describe sources of financial or non-financial support for the review, and the role of the funders or sponsors in the review. | pp.2,9,24 |
| Competing interests | 26 | Declare any competing interests of review authors. | p.24 |
| Availability of data, code and other materials | 27 | Report which of the following are publicly available and where they can be found: template data collection forms; data extracted from included studies; data used for all analyses; analytic code; any other materials used in the review. | p.24 |

**Table S2. Bibliographic search**

|  | **Concept** | **Search strings** |
| --- | --- | --- |
| 1 | General air pollution terms | "air adj3 pollut*" or "air adj3 quality" or "AQI" or "ambient air" or "outdoor air" or “aerosols” |
| 2 | Criteria air pollutants | "particulate matter" or "PM2?5" or "PM10" or "nitrogen dioxide" or "NO2" or "NOx" or "sul* oxid*" or "sul* dioxid*" or "SO2" or "ozone" or "O3" or "carbon monoxide" or "lead poison*" or "lead exposure" |
| 3 | Hazardous air pollutants | (("volatile organic compounds" or "VOCs" or "benzene" or "toluene" or "formaldehyde" or "polycyclic aromatic hydrocarbons" or "PAHs" or "benzo[a]pyrene" or "arsenic" or "cadmium" or "chromium" or "mercury" or "methylmercury" or "nickel" or "dioxins" or "furans" or "polychlorinated biphenyls" or "PCBs" or "persistent organic pollutants" or "POPs" or "hexachlorobenzene" or "HCB" or "pesticides" or "DDT" or "organochlorine pesticides" or "organophosphate pesticides" or "atmospheric pesticides" or "endocrine disrupting chemicals" or "EDCs") adj3 ("air pollution" or "airborne" or "atmospheric" or "ambient air" or "air quality" or "aerosols") |
| 4 | Emerging air pollutants | ("black carbon" or "brown carbon" or "microplastic*" or "nanoplastic*" or "ammonia" or "ultrafine adj2 partic*" or "UFPs") adj3 ("air pollution" or "airborne" or "atmospheric" or "ambient air" or "air quality" or "aerosols") |
| 5 | Sex ratio terms | ("sex ratio" or "male-to-female birth ratio" or "likelihood of giving birth to girls" or "likelihood of giving birth to boys" or "proportion of male births" or "proportion of female births" or “proportion of males” or “proportion of females” or “male birth probability” or “female birth probability” or “probability of male birth” or “probability of female birth” or “giving birth to male*” or “giving birth to female*” or "male birth rate" or "female birth rate" or "gender ratio at birth" or "birth ratio" or "sex distribution at birth" or "male-to-female ratio" or “male to female ratio” or “female-to-male ratio” or “female to male ratio” or “male birth* per female birth*” or “female birth* per male birth*” or “birth masculinity” or “birth femininity”) not animal not bovine not cat not dog not rat not mice not frog not fish not poultry not insect not parasit* not bee |
| 6 |  | 1 or 2 or 3 or 4 |
| 7 |  | 5 and 6 |
| 8 |  | remove duplicates from 7 |

The free text search terms (above) were used to identify studies in MEDLINE and Embase (tw,kf) and Global Health (ti,ab). Free text terms for air pollution were combined with the following MeSH fields in MEDLINE and Emtree in Embase.

MeSH MEDLINE: environmental pollution/ or air pollution/ or traffic-related pollution/ or environmental pollutants/ or air pollutants/ or exp particulate matter/

Emtree Embase: exp air pollution/ or traffic pollution/ or pollution/ or pollutant/ or exp air pollutant/ or exp particulate matter/

**Table S3. Adapted NIH Quality Assessment Tool for Observational Cohort and Cross-Sectional Studies (Risk of Bias)**

|  | **Criteria** | **Detail** |
| --- | --- | --- |
| 1 | Clear objective | Is the objective to evaluate the association between air pollution and SRB clearly defined? |
| 2 | Population defined | Is the source population clearly described, with eligibility criteria? |
| 3 | Representative sample | Was the study population representative of the broader population (e.g., national registry, full region)? |
| 4 | Temporal resolution of pregnancy exposure | Was air pollution exposure linked at a meaningful temporal level (e.g., gestational week, trimester, vs. whole pregnancy average)? |
| 5 | Exposure measurement validity | Was exposure assigned at the individual- or area-level? Were pollutants measured via validated methods? |
| 6 | Exposure precedes outcome | Was exposure to air pollution measured before the outcome occurred (birth)? |
| 7 | Valid outcome measure | Was the SRB measured using reliable data? |
| 8 | Confounding controlled for adequately | Were relevant confounders adjusted for (e.g., seasonality, SES, place-specific characteristics, maternal characteristics)? |
| 9 | Statistical methods appropriate | Were models appropriate for the design and data type? Were sensitivity or interaction analyses used? |
| 10 | Design specific strength | Did the study have any plausible claims to causality with its design? |

**Table S4. Risk of Bias Scores**

| **Study** | **Clear objective** | **Population defined** | **Representative sample** | **Temporal resolution of pregnancy exposure** | **Exposure measurement validity** | **Exposure precedes outcome** | **Valid outcome measure** | **Confounding controlled for adequately** | **Statistical methods appropriate** | **Design-specific strength** | **Overall**  **risk of bias**  **(/20)^1^** |
| --- | --- | --- | --- | --- | --- | --- | --- | --- | --- | --- | --- |
| Arima et al (2023) | 2 | 1 | 1 | 0 | 1 | 1 | 2 | 0 | 1 | 0 | **9** |
| Candela et al (2013) | 2 | 2 | 1 | 1 | 1 | 2 | 2 | 2 | 2 | 1 | **16** |
| Ghosh et al (2019) | 2 | 2 | 1 | 2 | 2 | 2 | 2 | 2 | 2 | 1 | **18** |
| Goin et al (2024) | 2 | 1 | 2 | 2 | 2 | 2 | 2 | 2 | 2 | 2 | **19** |
| Houdek et al (2020) | 0 | 0 | 1 | 0 | 1 | 0 | 2 | 1 | 1 | 0 | **6** |
| Lin et al (2006) | 2 | 2 | 1 | 0 | 1 | 0 | 2 | 1 | 1 | 1 | **11** |
| Lin et al (2015) | 2 | 2 | 0 | 2 | 1 | 2 | 2 | 2 | 2 | 1 | **16** |
| Miraglia et al (2013) | 2 | 1 | 1 | 0 | 1 | 0 | 2 | 0 | 0 | 0 | **7** |
| Sanders & Stoecker (2015) | 1 | 2 | 2 | 0 | 1 | 0 | 2 | 2 | 2 | 2 | **14** |
| Santoro et al (2016) | 2 | 2 | 0 | 0 | 1 | 0 | 2 | 1 | 1 | 0 | **9** |
| Tsai et al (2025) | 2 | 1 | 1 | 0 | 1 | 0 | 2 | 0 | 0 | 0 | **7** |
| ^1^  Risk of bias score out of (n/20); the higher the score, the lower the risk of bias. High risk: 0-10; medium risk: 11-15; low risk: 16-20. Study quality was appraised using an adapted version of the NIH Quality Assessment Tool for Observational Cohort and Cross-Sectional Studies (NHLBI 2014), modified to accommodate ecological, quasi-experimental, and time-series designs. | | | | | | | | | | | |
